# Psychosocial effects of Cutaneous Leishmaniasis on Patients in Northwestern Ethiopia: A qualitative study

**DOI:** 10.1101/2023.10.25.23297130

**Authors:** Helina Fikre, Yassin Mohammed Yesuf, Saba Atinafu, Tadele Mullaw, Deladergachew Dessie, Zemenay Mulugeta, Azeb Tadesse, Johan van Griensven, Saskia van Henten

## Abstract

Cutaneous Leishmaniasis (CL) is a serious skin disease prevalent in many countries around the world, and in Ethiopia, it presents with stigmatizing lesions sometimes compared to leprosy. However, little attention has been given to the psychosocial impact of CL on affected individuals.

To address this gap, we conducted a qualitative study using in-depth interviews with 13 CL patients (three patients with diffuse CL, three with localized CL, and seven with mucocutaneous leishmaniasisis).

Interpretative phenomenological analysis was used to analyze the qualitative data collected from the in-depth interviews. Our analysis revealed two themes: psychological burden, and social burden. As to psychosocial burden, patients reported experiencing anger, sadness, shame, fear, and hopelessness, along with feelings of inferiority.. Related to the social burden of CL, self-stigmatization, stigma, disrupted life, disability, community reaction, and lack of social support were identified as subthemes.

These findings highlight the overlooked psychosocial problems faced by Ethiopian CL patients and underscore the need for patient-oriented treatment packages. Policymakers and researchers should focus on creating community awareness on CL along with integrating psychological interventions.

## Background

Cutaneous leishmaniasis (CL) is a neglected tropical disease that causes patients to feel stigmatized and inferior in their daily lives(1). It is considered one of the most serious skin diseases in developing countries, where the common clinical presentations are severe and present with stigmatizing lesions that are sometimes compared with leprosy(1). Ethiopia is one of the ten countries with a high burden of CL, and *Leishmania aetiopica* is the most common cause of the disease in the country(2). This species is known to cause disfiguring lesions, including mucocutaneous leishmaniasis (MCL) and diffuse cutaneous leishmaniasis (DCL)(2,3). DCL is the most severe form of CL characterized as non-ulcerating papular, nodular, and plaque lesions involving most parts of the body(3). MCL is characterized by mucosal involvement (nasal, oral, pharynx, larynx)(2,3).

Studies have shown that patients with CL experience negative psychological impacts, such as anxiety, stress, depression, and low quality of life, which ultimately affect their economic productivity(4,5). Moreover, many individuals affected by CL tend to delay seeking treatment due to negative perceptions and attitudes towards modern treatment options and a lack of information about CL treatment, which results in more extensive skin lesions and pronounced stigma(6).

Although leishmaniasis-related stigma has been reported in several countries(1), cultural, religious, environmental, and geographical factors affect the perception of CL and its psychosocial effect. As such, results cannot be directly extrapolated from other settings. A study in Suriname has shown there is no significant stigma related with CL(7), and on the other hand a study from Morocco has shown that permanent CL scars lead to self-stigma and social stigma, which leads to a negative psychological effects in adolescent age group(8). This shows that not all patients with CL have the same experience. A study done in Ethiopia on quality of life (qol) of CL showed very poor health related quality of life (HRQoL) in almost half of the patients(9). Another study done on prevalence and stigma related with CL on patients suspected to have CL in northwest Ethiopia has showed stigma among the community was significant for CL cases(10).

In Ethiopia, the psychosocial impact of cutaneous leishmaniasis (CL) on patients has not been thoroughly explored. This qualitative study aimed to understand the psychological effects experienced by CL patients in northwest Ethiopia. By gathering and analyzing patients’ first-hand experiences, we can gain insights into their daily lives, beliefs, and values. This understanding is crucial for delivering patient-centered treatment that addresses not just their physical symptoms but also their social and psychological well-being, ultimately improving their overall quality of life.

## Methods

### Setting

The Leishmania Research and Treatment Center (LRTC) is a collaborative effort between the University of Gondar (UoG) and the Drugs for Neglected Diseases Initiative (DNDi) in northwest Ethiopia.

LRTC primarily focuses on reseach and treating visceral leishmaniasis (VL) but also provides care for cutaneous leishmaniasis (CL) patients from various endemic districts in the Amhara region. CL patients are referred to LRTC from peripheral health facilities or the dermatology departments of the UoG hospital. Diagnostic and treatment services at LRTC are provided at no cost.

### Population

From January to August 2021, all patients seeking care at the LRTC, UoG, diagnosed with CL (through clinical or parasitological confirmation) were considered for the study Inclusion criteria comprised of age above 18 and willingness to participate, while individuals with mental illness, acute disease, or existing chronic skin conditions were excluded.

Purposive sampling ensured varied representation in terms of gender, socioeconomic status, illness duration, and CL clinical features. Participant enrollment continued until data saturation.

### Interviews

Qualitative in-depth interviews were done to gain insight into the psychosocial experiences of CL patients. Qualitative in-depth interviews were conducted using open-ended questions, using the same interview guide for all patients. A physician trained in qualitative methods conducted the in-depth interviews, which were digitally recorded. On average, each interview lasted 30 minutes. Interviews were conducted in Amharic, the mother tongue of both patients and investigators, ensuring the accuracy of the transcripts.

### Data analysis

Qualitative data from in-depth interviews was analyzed using interpretative phenomenological analysis in Microsoft Excel. Transcripts were first transcribed in Amharic, then translated to English after themes were developed. The text was coded, clustered, categorized, and sub-categorized to identified meaning units. Themes and subthemes were constructed to interpret the latent meaning, ensuring trustworthiness through concurrent analysis and team discussion.

### Quality control

Trustworthiness of the study was ensured through various means. The researchers secured the confirmability of the research by maintaining the documentation at all stages of the research. The same interview guide was used for all the patients. The study was performed through teamwork and under the guidance and supervision of experts, which enhanced the dependability and confirmability of the data.

### Ethics

Ethical considerations were also taken into account. The study protocol and informed consent forms were approved by the Institutional Review Committee (IRB) of UoG. All the participants gave oral informed consent, and the data presented contains no identifying information.

## Results

A total of 13 patients were included in the study, which are described in table 1. Seven had MCL, three had DCL, and three had LCL. The mean and median age of the participants were 35 and 22.5 years old, respectively. Seven were male, and seven were female. Four were students, four were farmers, one was a priest, two were housewives, and three were unemployed. The duration of illness ranged from 1 year to 17 years.

**Table 1.** demography of participants, MCL-Mucocutaneous leishmaniasis, DCL-Diffused cutaneous leishmaniasis, LCL-Localized cutaneous leishmaniasis. The interview Id numbers are only known to reseach team

| **No** | **Age** | **Gender** | **Marriage** | **Occupation** | **Duration** | **Type of CL** |
| --- | --- | --- | --- | --- | --- | --- |
| 1 | 18-25 | Male | Single | Deacon | 1 year | MCL |
| 2 | 18 -25 | Male | Single | Unemployed | Unknown<br>(Long) | DCL |
| 3 | 50-55 | Male | Divorced | Painter | 7 years | DCL |
| 4 | 18-25 | Female | Single | Student | 1 year | LCL |
| 5 | 18-25 | Male | Single | Student | 3 years | LCL |
| 6 | 18-25 | Male | Single | Merchant | 1 year and<br>3 months | MCL |
| 7 | 18-25 | Male | Single | Student | More than<br>a year | LCL |
| 8 | 56-60 | Male | Married | Farmer | 2 years | MCL |
| 9 | 56-60 | Female | Married | House<br>wife/farmer | More than<br>2 years | MCL |
| 10 | 40-45 | Female | Married | Farmer | 9 month | MCL |
| 11 | 65-70 | Female | Widowed | House wife | >1year | MCL |
| 12 | 50-55 | Male | Not<br>married | Farmer | 3 year | MCL |
| 13 | 18-25 | Male | Single | Unemploye<br>d | 13 years | DCL |

Most of the patients needed time and probing questions to feel comfortable talking about the effect of the lesion on their daily life. Two themes emerged from the analysis: psychological burden, and social burden, each with several subthemes

### Psychological burden

#### Anger/sadness

Some described the unhappy feeling they had while others felt sorry for themselves. On the other hand, some of the patients mentioned the feeling of anger and frustration because they had to live with CL.

One of the respondents has stated, “*I used to feel sorry for the people who have CL in the past and I feel sad that God has brought this unto me*”. *interview 9*

*I have no words to describe how much having CL has affected me, it was difficult having this lesion for the last 2 years, I suffered a lot, I wish no beast will have this let not alone a person”. interview 8*

#### Regret

Some participants showed feelings of regret, specifically their delay in seeking treatment.

A female respondent said “*I postponed seeking treatment because I had to be at home to take care of my babies and the days passed without getting treatment, but now I regret not coming earlier”. interview 10*

#### Fear of consequences

Fear is a typical psychological state reported by patients. Most patients fear CL might ruin and disfigure their faces and/or disable them for good.

Here a young male DCL patient said, “*I fear the lesion might ruin my body worse than it already has, and I may not survive”. interview 2*

In addition, most participant has stated that they fear the complication of the lesion might affect their other body and daily function.

an older woman is a good example here. She said: “*I fear if the lesion stays longer it might ruin my face, sometimes I fear I am having headache and eye pain is due to the lesion”. interview 11*

#### Feeling inferior

The majority of CL patients not only expressed their feelings of inferiority but also explained why they had those feelings. A good example here is an teen male respondent

“*It feels so bad, when it suddenly appeared on my face, the people who ask me questions are very annoying. It’s hard to always cover my face with a scarf whenever I go to school, this makes one feel inferior*.*” interview 4*

#### Shame

The majority of the respondents mentioned that they are ashamed of their illness as the lesion will make them feel they are different from others. They also feel ashamed because of the disfiguring nature of their CL and due to the comments they get from the community.

One of the participants emphasized, “*I feel ashamed because of how different I look, so I don’t feel comfortable mixing with others unless I have to*”. *interview 4*

#### Covering their lesions

Almost all patients covered their lesion because they feel uncomfortable walking around with the lesion exposed and being stared at. They also felt how others might judge them for acquiring this lesion. And in some areas, it is believed that patients have to cover the lesion so that it will not get worse.

One of the participant stated, *“I usually cover up my face whenever I go out the home but I won’t cover it in my house, plus it is also believed the lesion will worsen if it is seen by others”. interview 10*

#### Hopelessness

Hopelessness was observed in a small number of CL patients. These patients felt hopeless after a lengthy treatment and recurring lesion after being treated. One respondent who had lived with the lesion for many years stated.

*“Since the lesion never got cured but comes back every time, I have lost hope, now my brain is not thinking properly”. interview 1*

### The social burden of CL

#### Disrupted life

For more than half of the respondents, their earlier social relationships got ruined because of their disease, whereas some stopped their social interactions completely. For instance, a male patient in his twentys said the following:

*“I will not go whatever it is. In the early days, <u>I used to go with my relative</u> to my parent’s “medanialem maheber” [a religious gathering on the 27th day of the month]. But now I quit it. My friends used to invite me to weddings and family gathering but I never go. I won’t leave the house in the first place. I didn’t want to mix with people in general*.*” interview 13*

Most of the patients were unable to continue with their previous activity and work because of the lesion. Some have said working in the field, and being exposed to sunlight might worsen the lesion therefore; they would rather stay indoors.

“*It is believed that being exposed to the sun will worsen the lesion, therefore; it had limited me from working on farming”. interview 9*

Another participants replied “ *Because of the lesion first of all I can’t work, I can’t have fun, I am young but I can’t even play football with my friends because the lesion bleeds with the slightest touch”. interview 5*

Few patients were worried the lesion worsen during sex, and they don’t feel comfortable having sex with their partners. One patient refuses to get married because he feels hopeless and inferior due to his condition. Some said they are students and young therefore they don’t worry much about the lesion affecting their love life.

One respondant said *“I had a girlfriend before the lesion gets this extensive, she lives abroad, she always wants to settle but I don’t want to get married being this sick and disfigured”. interview 13*

Another respondant said *“ I fear the lesion worsens during sex, I still have sex but I am always in doubt”. interview 12*

#### Self-stigmatization

The majority of the patients distance themselves from their social circles and occasions like weddings or funerals unless they really have to go. They would rather stay at home.

Further scrutiny of the interview extracts depict that perceived and enacted discrimination from others in the community are behind patients’ self-discrimination.

One respondent said *“it will take me a month to tell you how much the lesion affected me, I spend my days on the field with my cattle so no one sees me. People think I was always like this, they don’t know me when I was well. Sometimes when I want to have a soft drink I used bajaj (three wheel transportation) to go to a café and sit inside where no one sees me”. interview 8* Some participants have also stated even though they want to attend church service they are not able to because some say, it is a sin to go closer to church with their lesion and they fear other people’s opinions.

One participant said “*I stopped going to church but if I have to I will go early in the morning when no one is around, I really gave up then”. interview 7*.

#### Disability

Few patients were disabled because of the disfiguring nature of the lesion. They said it has limited them from doing things because of the lesion, even from routine activities like eating.

This was well explained by one of the participants,

*I was not able to eat, drink, and talk as I used to, I only took a meal with milk, because I couldn’t chew and swallow properly. Even then some of the meals and drinks will pour out of my mouth as I was not able to close my lips. I had many sleepless nights because of the pain over my lips, In the last two weeks it becomes so painful that I used to pour water to get some relief”. interview 8*

A young participant also stated “*I feel congested since the lesion has cut off my nose. And because I lost flexibility of both my hands I have stopped working altogether since last year”. interview 2*

#### Community reaction

According to patients, their friends and family treat them with empathy and accommodation. However, their wider circle of acquaintances, such as colleagues and classmates, treat them differently due to their lesion, often staring and mocking them.

One patient said, “ *Before coming to Gondar to seek treatment a lot of people who don’t know me used to be very scared when they see me, and especially kids used to run away from me because they were scared of me”. interview 4*

Almost all patients are saddened by the comments people give them, especially people they do not know whom they found on the road, in cafés, and in church.

A male participant said “ *When there is a special occasion I go to my neighbors or to the market also if there is a zeker (especially church ceremony) I will go to church but I won’t eat or drink because I am being judged for even going, people tell me to stay away from the church and stay at home”. interview 10*

Another one had stated *“I get angry when people comment and ask about my lesion”. interview 7*

#### Stigma

Most patients experienced stigma from people around themwhich was expressed in the form of direct insults and mistreatment.

A male participant stated *“ People sometimes smirk at me when the see me”. interview 6* Another female respondant said *“People stare at me and sometimes they insult me as if I brought this on myself”. interview 10*

Few were also being isolated for fear of being exposed to the infection as is thought to be contagious. They said they were being told to eat, walk and sit in isolation. This was well described by one participant.

*“When I was at school my classmates fear it is contagious so they isolate me, and they don’t want to walk with me, this make me feel stigmatized and ashamed”. interview 2*

#### Social support

Throughout their journey of seeking help and receiving treatment, patients were provided with significant support from various individuals, including priests and teachers.

This support includes financial assistance for transportation and accommodation, suggestions that strongly encourage treatment, and valuable guidance on where to seek both modern and traditional treatments.

There was one patient who said *“ my family has marriam mahiber “religious social gathering among close friends and her friends managed to raise 2000 Ethiopian birr and send me for treatment”. interview 13*

Another respondant said *“when I go to school my teacher always urgdt me to get treatment”. interview 4*

## Discussion

Our results her show the depth of psychological and social burden patients endure when living with CL. Our participants experienced anger, sadness, and also feelings of inferiority, similar to another studydone in Yemen that found that loss of self-esteem and feelings of inferiority were associated with CL (1).

Though most patients wished to get cured and go back to their old life they still fear the lesion might ruin their face permanently or disable them, or the scar may persist even after being cured. This aligns with a study in Morroco that has observed adolescents living in the region affected by CL had a psychological impact,also stating CL causes more psychiatric problems as it leaves a permanent scar on the affected area(8).

Some participants felt hopeless and did not see why they are still alive, this was also shown in a study done in Morocco on adolescents who reported a large range of negative psychosocial effects, ranging from slight shame to suicidal thoughts(8).

Although we did not assess our patients for depression, most of the patients were sad, angry, hopeless, and felt inferior because of CL, which may lead to developing depressive symptoms if not intervened earlier. A study where depressive symptoms were compared between children and adolescents with CL and health control in Turky showed depressive symptoms were higher in cases with CL(11). Our findings also aligned with available evidence from a systemic review which has estimated that 70% of patients with both active and inactive CL will experience some degree of psychological morbidity(12).

In this study, patients experienced significant social burdens and disruptions in their lives due to their condition. They feared insults and hurtful comments, refrained from social gatherings, and some were even physically disabled.

Additionally, the societal belief that their illness was a sin caused sadness and self-stigmatization, leading them to isolate themselves at home. In contrast, a study in Suriname found negative reactions due to CL are basically only for a small group of patients who endured more severe forms of CL(7). As most of our patients had more severe forms of disease, which is typical for CLpatients who visit the hospital in Ethiopia, it is hard to know to what extent these findings also apply for less severe lesion types.

Almost all of our patients also felt stigma from the community, some were told to isolate themselves from gatherings, church or to eat separately because its thought to be contagious. A study done in Suriname showed that the nature of stigma that patients experience are associated with certain general fears and anticipation of rejection, rather than the actual rejection that patient had experienced from society(7). Stigma in Ethiopia, specifically in the Amhara region where all of our participants lived, could be due to misconceptions on modes of transmission, as a study has shown that the community believes CL is transmitted by touch(6). Additionally, religion may contribute to some misconceptions, as several orthodox Christian patients wrongly perceived CL as caused by a curse or God’s punishment for one’s sinful deeds(6).

Our study revealed that patients were unable to maintain their previous employment, fearing that exposure to the sun during field work or being seen by others worsened their condition. This resulted in reduced income and limited access to healthcare. Additionally, religious misconceptions and beliefs had a profound psychological and social effect on the patients. This valuable information can serve as a foundation for community interventions aimed at addressing these issues. A review of community interventions found that education and innovative approaches targeting stigma, financial barriers, and access to healthcare can improve knowledge, attitudes, and practices in communities.(13).

Our study has several limitations. Findings reported here may not be representative as this was a hospital-based study. As such, most of our participants were affected by severe forms of CL as the majority seeks treatment very late. Therefore, the findings may not reflect the psychosocial burden in patients with smaller lesions and short illness duration.

## Conclusion

Our study brings attention to the psychosocial effects of CL based on patients’ experiences. Negative effects were observed as patients isolated themselves due to community and self-stigma. To address this, we recommend practitioners to raise awareness, improve access to diagnosis and treatment, integrate psychological support, and provide reconstruction surgery for CL-related disabilities.

## Data Availability

All data produced in the present study are available upon reasonable request to the authors

## Acknowledgment

We would like to thank the study team. We also acknowledge the patients who have agreed to share their information irrespective of their condition. We also thank the Institute of Tropical Medicine for supporting the study with expertise and funding.

## Research ethics and patient consent

We have obtained ethical approval from the University of Gondar institutional review board. Consent was obtained from all study participants before being enrolled in the study.

## References

1. Al-Kamel MA. Stigmata in cutaneous leishmaniasis: Historical and new evidence-based concepts. Our Dermatology Online [Internet]. 2017;8(1):81–90. Available from: http://www.odermatol.com/issue-in-html/2017-1-21-leishmaniasis/

2. Ethiopian ministry of health. GUIDELINE FOR DIAGNOSIS AND PREVENTION OF LEISHMANIASIS IN ETHIOPIA. Addis Abeba; 2013. 2–3 p.

3. Goto H, Lauletta Lindoso JA. Cutaneous and Mucocutaneous Leishmaniasis. Infect Dis Clin North Am. 2012;26(2):293–307.

4. Yan CH, Yao, Hong S, Fung, Lui Y Ji, et al. Dynamic Balancing in Illness Coping: An Interpretative Phenomenological Analysis on the Lived Experience of Chinese Patients with Psoriasis. Heal Sci J [Internet]. 2017;11(4):1–10. Available from: http://www.hsj.gr/medicine/dynamic-balancing-in-illness-coping-an-interpretative-phenomenological-analysis-on-the-lived-experience-of-chinese-patients-with-p.php?aid=20035

5. Behrooze Vares MD1, Mina Mohseni2, Amireh Heshmatkhah MD1, Saeideh Farjzadeh MD1 3, Simin MD, Shamsi-Meymandi 4, Zahra Rahnama MD4, Leila Reghabatpour MD4 OFM. Quality of Life in Patients with Cutaneous Leishmaniasis. 2013;16(8):474–7. Available from: http://www.ams.ac.ir/AIM/NEWPUB/13/16/8/008.pdf

6. Tamiru HF, Mashalla YJ, Mohammed R, Tshweneagae GT. Cutaneous leishmaniasis a neglected tropical disease□: community knowledge, attitude and practices in an endemic area, Northwest Ethiopia. 2019;1–10.

7. Ricardo V. P. F. Hu, Sahienshadebie Ramdas, Pythia Nieuwkerk, Ria Reis, Rudy F. M. Lai A Fat, Henry J. C. de Vries, Henk D. F. H. Schallig. Body location of “ New World “ cutaneous leishmaniasis lesions and its impact on the quality of life of patients in Suriname. 2020;(Cl):1–12. Available from: 10.1371/journal.pntd.0008759

8. Bennis I, Thys S, Filali H, De Brouwere V, Sahibi H, Boelaert M. Psychosocial impact of scars due to cutaneous leishmaniasis on high school students in Errachidia province, Morocco. Infect Dis Poverty. 2017;6(1):1–8.

9. Shimelis Doni, Kidist Yeneneh, Yohannes Hailemichael, Mikyas Gebremichael, Sophie Skarbek, Samuel Ayele, Abay Woday, Saba Lambert SL, Walker EG. Health-Related Quality of Life of Adults with Cutaneous Leishmaniasis at ALERT Hospital, Addis Ababa, Ethiopia. 2023;1–15.

10. Lamesgen A. Prevalence, Level of Stigma, and Associated Factors of Cutaneous Leishmaniasis Among Suspected Peoples Living in Dega Damot District, [Internet]. 2022. Available from: http://ir.bdu.edu.et/handle/123456789/14942

11. Turan E, Kandemir H, Yeşilova Y, Ekinci S, Tanrikulu O, Kandemir SB, et al. Assessment of psychiatric morbidity and quality of life in children and adolescents with cutaneous leishmaniasis and their parents. Postep Dermatologii i Alergol. 2015;32(5):344–8.

12. Baileyid F, Mondragon-Shem K, Haines LR, Olabi A, Alorfi A, Ruiz-Postigo JA, et al. Cutaneous leishmaniasis and co-morbid major depressive disorder: A systematic review with burden estimates. PLoS Negl Trop Dis. 2019;13(2):1–22.

13. Polidano K, Wenning B, Ruiz-Cadavid A, Dawaishan B, Panchal J, Gunasekara S, et al. Community-Based Interventions for the Prevention and Control of Cutaneous Leishmaniasis: A Systematic Review. Soc Sci. 2022;11(10).

